# THE EXPERIENCE OF UK BLADDER CANCER PATIENTS DURING THE COVID-19 PANDEMIC: A SURVEY-BASED SNAPSHOT

**DOI:** 10.1101/2020.06.27.20140582

**Authors:** Sarah Spencer-Bowdage, Jeannie Rigby, Jackie O’Kelly, Phil Kelly, Mark Page, Caroline Raw, Paula Allchorne, Peter Harper, Jeremy Crew, Roger Kockelbergh, Allen Knight, Richard T Bryan

**Author notes:** **Correspondence to:** Mr Richard T Bryan, The Bladder Cancer Group, Institute of Cancer & Genomic Sciences, University of Birmingham, Edgbaston, Birmingham, B15 2TT, UK. **Disclosures:** RT Bryan has contributed to advisory boards for Olympus Medical Systems & Janssen, and undertakes research funded by UroGen Pharma.

## Abstract

The Covid-19 pandemic has placed unprecedented strain on healthcare systems worldwide. Within this context, UK cancer services have undergone significant disruption to create capacity for the National Health Service. As a charity that endeavours to support bladder cancer (BC) patients and improve outcomes, Action Bladder Cancer UK (ABCUK) designed and administered a *SurveyMonkey* survey to investigate the prevalence of such disruption for BC patients. From 22^nd^ April to 18^th^ June 2020, 142 BC patients responded. Across all patient groups, 46.8% of patients described disruption to their treatment or follow-up. For non-muscle-invasive BC (NMIBC) patients, disruptions included postponement of: initial transurethral resection of bladder tumour (TURBT) (33.3%), subsequent TURBT (40.0%), and surveillance cystoscopy (58.1%). For NMIBC patients undergoing intravesical therapy, 68.4% experienced treatment postponements or curtailments. For muscle-invasive BC patients, 57.1% had experienced postponement of cystectomy and 14.3% had been changed from cystectomy to radiotherapy. Half of patients undergoing systemic chemotherapy also experienced disruption. Despite the survey’s limitations, we have demonstrated considerable disruption to the care of BC patients during the UK Covid-19 pandemic. To avoid a repeat, the UK BC community should define effective contingent ways of working ready for a possible ‘second wave’ of Covid-19, or any other such threat.

## BRIEF CORRESPONDENCE

The Covid-19 pandemic has placed unprecedented strain on healthcare systems worldwide with the requirement to treat large influxes of infected patients, many of whom require respiratory support. Understandably, healthcare systems have had to redirect resources and redeploy staff away from routine diagnostic, treatment and follow-up services. The UK’s National Health Service (NHS) is no different, with staff demonstrating remarkable resilience and flexibility whilst themselves suffering Covid-19 morbidity and mortality. Within this context, cancer services have undergone significant disruption to create the capacity for the NHS to tackle the pandemic.

In the UK there are over 10,000 new diagnoses of bladder cancer (BC) per year with at least 50,000 patients undergoing long-term follow-up or surveillance, the majority of whom have life-limiting or life-threatening disease. Despite rapid and comprehensive recommendations on the prioritisation of specific uro-oncology patient subgroups from the British Association of Urological Surgeons (BAUS), the European Association of Urology (EAU), and other sources [1-4], many patients will have inevitably experienced considerable disruption to their treatment and follow-up regimens either due to service pauses or safety concerns in the pandemic environment [5-7]. In this regard, and as a charity that endeavours to support bladder cancer patients and improve outcomes, Action Bladder Cancer UK (ABCUK, Tetbury, UK) designed and administered a survey to investigate the prevalence of such disruption.

Utilising the *SurveyMonkey* (San Mateo, CA 94403, USA) platform, and following several reviews and re-iterations by charity trustees, the survey was launched on 22^nd^ April 2020 (c.10 days following the peak of UK Covid-19 cases). The questionnaire and accompanying CHERRIES checklist [8] can be viewed in the Supplementary Files. UBC patients were directed to the survey via the ABCUK website (http:/actionbladdercanceruk.org/) and social media platforms. From inception to censor date (18^th^ June 2020), 142 bladder cancer (BC) patients had responded. Summary responses from the survey can be viewed in Supplementary Data.

Over 95% of respondents lived in England, although there was geographical reach from all of the UK, including rural areas and major conurbations such as West Yorkshire, Greater Manchester and Greater London. Almost 80% of respondents were aged 60 years or older and over 72% were male, reflecting the age and gender distribution of BC in the UK population [9]. Almost 72% of respondents were non-muscle-invasive bladder cancer (NMIBC, stages Ta/T1/Tis) patients, 25% were muscle-invasive bladder cancer (MIBC, stages T2+) patients, and 3% had a diagnosis of advanced or metastatic disease.

Across all groups (139/142 responses), 46.8% of patients described disruption to their treatment or follow-up (delays, postponements, or treatment cancellations or curtailments); 34.5% of patients indicated no change, with treatment and follow-up proceeding as normal. The majority of the remaining 18.7% of patients were scheduled for follow-up several months in the future and had not yet been informed of any changes.

In patients who described disruption, 51.2% had received a telephone call to inform them, 26.8% had received a letter, 2.4% had received a text message, and 19.5% had contacted the hospital themselves. Perhaps understandably, 42 (33.3%) of respondents reported that the Covid-19 crisis had made it more difficult to communicate with their hospital urology team.

Six respondents were awaiting their initial transurethral resection of bladder tumour (TURBT); for 2 of these patients (33.3%), TURBT had been delayed or postponed. Fifteen patients were awaiting a subsequent TURBT, either for re-resection or to treat recurrence; for 6 of these patients (40.0%), TURBT had been delayed or postponed. Seventy-four NMIBC patients described being under cystoscopic surveillance; for 43 of these patients (58.1%) surveillance had been delayed or postponed. Of 57 NMIBC patients undergoing courses of intravesical therapy, 39 (68.4%) described delays, postponements or curtailments in treatment.

Fourteen patients were awaiting cystectomy, of which 8 patients (57.1%) had been notified of a postponement in their surgery, and 2 patients (14.3%) had been notified of cancellation of their surgery; these latter 2 patients had their treatment plan changed from cystectomy to radiotherapy. There were no patients whose treatment plan had changed from radiotherapy to cystectomy.

Eight patients described undergoing treatment regimens for locally-advanced or metastatic disease (adjuvant chemotherapy following cystectomy or radiotherapy, or chemotherapy only); 4 patients (50%) described disruption in the administration of chemotherapy.

Regarding the pandemic itself, 64 respondents (46.7%) had been advised to shield, and the majority of the remainder felt that they should have been advised to shield and decided to shield anyway. Almost 77% of patients expressed some concern about attending hospital for their treatment and follow-up appointments, and around 70% described that safety precautions for themselves and for staff would make them feel safer when attending hospital.

Clearly, a survey of this nature has a number of limitations. Although the patient demographics accurately reflect those of the UK UBC population, this remains a small study utilising an unvalidated questionnaire. The survey was more likely to be completed by existing BC patients who are already engaged with ABCUK, rather than newly-diagnosed patients who may not yet be aware of the work of the charity. Furthermore, local or regional patterns have not been comprehensively captured comprehensively, and more nuanced responses to the survey may have been obtained via telephone interview. Nevertheless, we have demonstrated the occurrence of considerable disruption to the care of bladder cancer patients in the UK during the Covid-19 pandemic.

UK BC patients are not alone in experiencing disruption to their care during the pandemic [2;6;7;10]. Some of these disruptions will have been justified in order to protect patients from Covid-19 itself or from additional complications of treatments in the environment of the pandemic [2;5;10;11]; yet, much disruption will have directly resulted from the redeployment of healthcare services to tackle the pandemic. The need for redeployment will not have been uniform, but it is likely that healthcare services in large population centres were more vulnerable. However, our survey appears to demonstrate that both MIBC and NMIBC patients have been equally affected by delays, postponements and cancellations during the Covid-19 pandemic. Hence, despite a plethora of recommendations from a number of sources outlining reasonable patient prioritisation strategies [1;3;4;10;12], these strategies may not have been developed and circulated quickly enough or enacted rapidly enough (or were unable to be actioned logistically) at or around the peak of the UK pandemic to have made a perceivable difference to patients themselves. Given the overwhelming nature of the pandemic on the whole of society, this is understandable. However, it is critical that the BC clinical and academic community maintains an ‘institutional memory’ should similar circumstances ensue in the future, either in the form of a ‘second wave’ of Covid-19, or as a separate threat; now is the time to plan contingent ways of working should either scenario become reality [13], and considerable evidence is available to inform such strategies (e.g. https://www.europeanurology.com/covid-19-resource, https://www.bjuinternational.com/bjui-blog/covid-19-collection-of-urology-papers/). Notwithstanding, the true success of any such strategy in the cancer setting can only be appropriately assessed several years downstream. We should also be aware that new suspected cancer referrals have also dramatically reduced during the pandemic [14], and so there is a long road to recovery ahead and none more so than for bladder cancer patients.

## Data Availability

Requests to access the data referred to in the manuscript can be made to Action Bladder Cancer UK or the corresponding author. Such requests will be reviewed by the charity Trustee Board in the context of the applicant's research objectives and pertinent regulatory permissions.

